# Spirometry performance quality and lung function pattern during pregnancy; should testing conditions and interpretation criteria be re-evaluated?

**DOI:** 10.1101/2023.06.02.23290897

**Authors:** Jacktan J Ruhighira, Fredirick L Mashili, Alexander M Tungu, Stephen Kibusi

**Affiliations:** Department of Physiology, University of Dodoma; Department of Physiology, Muhimbili University of Health and Allied Sciences; Department of Public Health, University of Dodoma

## Abstract

**Introduction:** Despite the prevalence of respiratory disorders in the gravid state, and the crucial role of spirometry in respiratory medicine, its utility for assessing lung function during pregnancy remains infrequent. Putative reasons for this include reservations regarding spirometry performance and its potential influence on test outcomes, although the literature documenting such concerns is insufficient. This study sought to evaluate whether variations in spirometry test performance could impact the diagnosis of pulmonary function patterns throughout gestation.

**Methods:** We used spirometry data from a cross-sectional study of 120 pregnant and 114 non-pregnant women who underwent spirometry with uniform instructions given to all subjects. Data were subjected to chi-square testing and subsequently evaluated through logistic regression analysis.

**Results:** The acceptable performance rate among pregnant participants was 77.3%, with the most common quality grade being C (37.5%). Pregnant individuals exhibited 2.1 times the odds of achieving a B grade (p=0.037, 95% CI=1.0-4.2) and 4.1 times the odds of achieving an F grade (p=0.02, 95% CI=1.6-9.9) instead of an A grade. Additionally, they manifested 2.9 times the odds of generating unsatisfactory performance (p=0.007, 95% CI=1.3-6.1) compared to non-pregnant participants. Also, pregnant participants displayed 2.5 times the odds of exhibiting a restrictive pattern (p=0.021, 95% CI=1.1-5.7); but pattern classification was not associated with quality grades.

**Conclusion:** Despite the higher likelihood of suboptimal spirometry quality, the observed pattern classification remains as expected physiologically, suggesting that spirometry is still a valid tool for assessing lung function in pregnancy.

## Introduction

Spirometry is a cost-effective and commonly used noninvasive lung function test. It assesses lung function by measuring the amount of air and speed of forceful exhalation (1). Despite its importance in respiratory medicine, spirometry is not usually used to assess lung function during pregnancy, although respiratory conditions are common in pregnancy (2). One of the reasons could be worrying about performance during pregnancy and its potential influence on test results. While this concern could hold, the documentation and evidence supporting it are insufficient.

The spirometry test must meet quality criteria to be useful in respiratory medicine. The American Thoracic Society/European Respiratory Society (ATS/ERS) has established spirometry acceptability and reproducibility criteria. The acceptable session has to begin with maximal inhalation, followed by an abrupt, most forceful exhalation without hesitation and continue smoothly without interruption by inhalation or cough for at least six seconds (3). The reproducibility is defined by the difference between the two largest forced vital capacities (FVC) and forced expiratory volumes in the first second (FEV1) (4).

Depending on quality and reproducibility, a quality control grade is assigned to the spirometry test at the end of the session. The criteria for performance grading (5) are described in Table 1. Grades A-C are considered clinically important and acceptable to interpret spirogram lung function patterns (6). There is a lack of studies to assess quality control grades during pregnancy which could probably determine which grades are clinically meaningful during this period.

**Table 1:** Spirometry performance quality control grades (5)

| Quality Control Grade | Number of Acceptable Manoeuvres | Repeatability (ml) |
| --- | --- | --- |
| A | 3 | <150 |
| B | 3 | 150-200 |
| C | 2 | 150-200 |
| D | 2 | >200 |
| E | 1 | NA |
| F | 0 | NA |

Even in non-gravid states, errors during spirometry such as submaximal inhalation, hesitations, coughing during the manoeuvre, early termination, and variable effort are common (7). With massive adaptations to pregnancy, spirometry can be expected to be even harder. Also, although spirometry values may remain in the normal range, they are lowered during pregnancy (8) which can be critical for those who had a borderline pre-pregnancy respiratory function. In the current analysis, we aimed to compare spirometry test performance and lung function patterns among pregnant and non-pregnant women and see if they were associated. Based on the normal physiology of pregnancy (9), we hypothesized that restrictive spirometry patterns would be more common in pregnant than in non-pregnant women, regardless of spirometry performance quality.

## Materials and Methods

### Study design and participants

We used data obtained from a cross-sectional spirometry study involving pregnant and non-pregnant women (8). We included 120 pregnant women randomly recruited from Mnazi Mmoja Hospital in Dares Salaam, Tanzania. Included pregnant participants were aged 18-35, had a 6-36 weeks singleton pregnancy, and booked their first antenatal visit during their first trimester. Ages under 18 were excluded due to presumed immature reproductive systems, while those above 35 were left out to exclude the effects of pregnancy in advanced age. The first five weeks were excluded due to the uncertainty of pregnancy diagnosis, while term pregnancy was excluded because of the increased risk associated with intrabdominal pressure during spirometry manoeuvre. Also, 114 non-pregnant participants were consecutively recruited from the Muhimbili University of Health Sciences (MUHAS) if they had similar characteristics except for being pregnant. Non-pregnant women who were pregnant in the preceding 42 days were excluded for possible effects of a previous pregnancy. Potential participants who had any contraindication for spirometry were excluded from the study. Detailed information on recruitment, inclusion, and exclusion criteria has been previously described (8).

### Spirometry test procedure and data collection process

Spirometry was done in a seating position using EasyOne® (ndd Medical Technologies, Zurich, Switzerland) digital spirometer set to diagnostic mode from May 10 to July 27, 2021. Participants were given similar instructions by the same technician adapted from the ATS/ERS 2019 spirometry standardization update (4) and American National Health and Nutrition Examination Survey (NHANES) 2011 manual (10). Everyone observed hand hygiene and kept a distance without facing each other directly. More details on spirometry procedures have been previously described (8).

### Variable measurements

The current analysis’s main outcome variables were spirometry test session quality control grade, acceptability, and lung function pattern. Each session was graded by the spirometer, and those graded A-C were considered acceptable while those graded D-F were regarded as unacceptable.

Only acceptable sessions were included in the analysis of the pattern. The definition of lung function pattern by spirometry (10) is described in Table 2.

**Table 2:** Definition of lung function pattern by spirometry (10)

| Lung Function Pattern | FEV1/FVC | FEV1% | FVC% |
| --- | --- | --- | --- |
| Normal | >70% | >80% | >80% |
| Obstructive | <70% | <80% | >80% |
| Restrictive | >70% | <80% | <80% |
| Mixed | <70% | <80% | <80% |

Exposure variables were participant characteristics; age, pregnancy status, body weight, height, parity, and gestational age. Age and parity were obtained by participant self-reporting. Height and body weight were measured using Seca® (Seca GmbH & Co. KG, Hamburg, Germany) scales. Weight was further categorized into body mass index (BMI) groups, but BMI in pregnancy was obtained using weight recorded in the first trimester instead of unavailable pre-pregnancy weight. Gestational age was obtained from an obstetric ultrasound done in the first trimester.

### Data analysis

Data were coded, entered, and analyzed using SPSS version 26 software. Participant characteristics were described using frequencies and percentages. The respective characteristic differences between pregnant and non-pregnant participants were analyzed by the chi-square test. The difference between pregnant and non-pregnant spirometry performance acceptability, quality control grades, and lung function patterns were preliminary analyzed by the chi-square test and further by logistic regression. Since the two groups under comparison differed in body weight and it was found to have an association with quality control grades, the difference was adjusted for body weight using ordinal regression analysis.

### Ethical consideration

The study was cleared by MUHAS’s ethical review board, and permission to conduct the study was sought from administrative authorities. A pre-arranged written informed consent was provided and signed by all participants. All test procedures were carried out per Helsinki’s declaration. Although spirometry is considered safe during pregnancy (11), safety precautions were taken, including taking a test in a seating position. Potential participants for whom spirometry was contraindicated were excluded from the study. Participants diagnosed with dysfunctional patterns were referred for further evaluation.

## Results

### Characteristics of study participants

This analysis included 234 participants, among whom 120 were pregnant and 114 were non-pregnant women (Table 3). The majority of participants were aged between 20-24 (38.1%), had normal body weight (47.5%), had a height of 150-159 cm (51.3), and had never given birth (48.6%). Most pregnant participants were in their third trimester between 28-35 weeks of gestation (44.1%). The pregnant and non-pregnant participants differed by age (p-value <0.001), body weight (p-value <0.043), and parity (p-value <0.001).

**Table 3:** Characteristics of Study Participants (n=234)

| Characteristic | | Pregnant<br>Frequency (%) | Non-pregnant<br>Frequency (%) | $p$-value* |
| --- | --- | --- | --- | --- |
| *Age (years)* | 18-19 | 3 (2.5) | 12 (10.5) | <0.001 |
|  | 20-24 | 32 (26.7) | 58 (50.9) |  |
|  | 25-29 | 36 (30) | 20 (17.5) |  |
|  | 30-35 | 49 (40.8) | 24 (21.1) |  |
| *Body weight* | Underweight | 6 (5) | 10 (8.8) | 0.043 |
|  | Normal weight | 50 (41.7) | 61 (53.5) |  |
|  | Overweight | 39 (32.5) | 20 (17.5) |  |
|  | Obese | 25 (20.8) | 23 (20.2) |  |
| *Body height* | 135-139 | 1 (0.8) | 0 (0) | 0.147 |
|  | 140-149 | 11 (9.2) | 12 (10.5) |  |
|  | 150-159 | 70 (58.3) | 50 (43.9) |  |
|  | 160-169 | 37 (30.8) | 47 (41.2) |  |
|  | 170+ | 1 (0.8) | 5 (4.4) |  |
| *Parity* | 0 | 42 (35) | 71 (62.3) | <0.001 |
|  | 1 | 41 (34.2) | 17 (14.9) |  |
|  | 2 | 19 (15.8) | 13 (11.4) |  |
|  | 3 | 14 (11.7) | 6 (5.3) |  |
|  | 4 | 4 (3.3) | 7 (6.1) |  |
| *Gestational Age* | 1st trimester | 11 |  |  |
|  | 2nd trimester | 34 |  |  |
|  | 3rd trimester | 75 |  |  |
|  | <20 weeks | 26 |  |  |
|  | 20-28 weeks | 41 |  |  |
|  | 28-35 weeks | 53 |  |  |

\**p*-value for the characteristic difference between pregnant and non-pregnant participants

### Spirometry performance during pregnancy

Acceptable performance was 77.3% (Figure 1) among pregnant participants, with the majority achieving grade C (37.5%), while it was 90.4% amongst non-pregnant participants, with the majority achieving grade B (30.7%). With reference to grade A, pregnant participants had 2.1 times the odds of achieving a B grade (p=0.037, 95% CI=1.0-4.2) and 4.1 times the odds of achieving an F (p=0.02, 95% CI=1.6-9.9) than their non-pregnant counterparts. When adjusted for body weight, pregnant participants had 3.8 times the odds of achieving a grade below A (p=0.001, 95% CI=1.7-8.2) than non-pregnant participants as weight increased. Moreover, pregnant participants had 2.9 times the odds of producing unacceptable performance (p=0.007, 95% CI=1.3-6.1) than non-pregnant participants.

**Figure 1:**
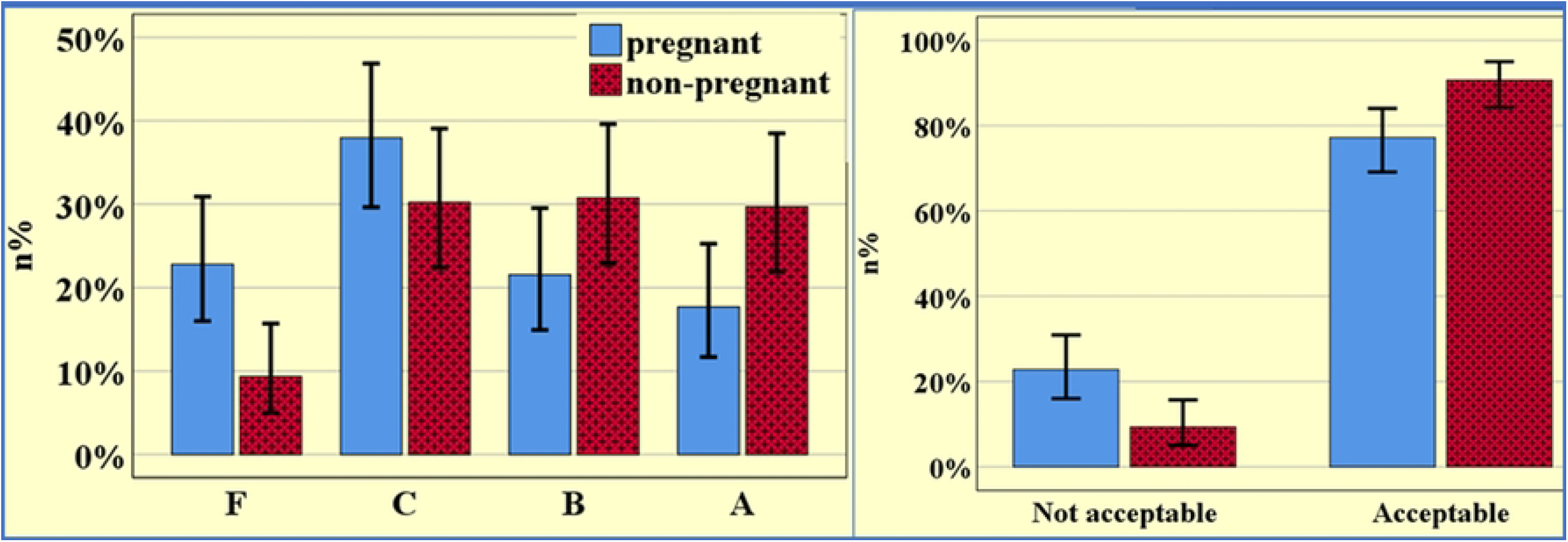
Spirometry performance of pregnant and non-pregnant participants (n=234, Error bar=95% CI)

**Figure 2:**
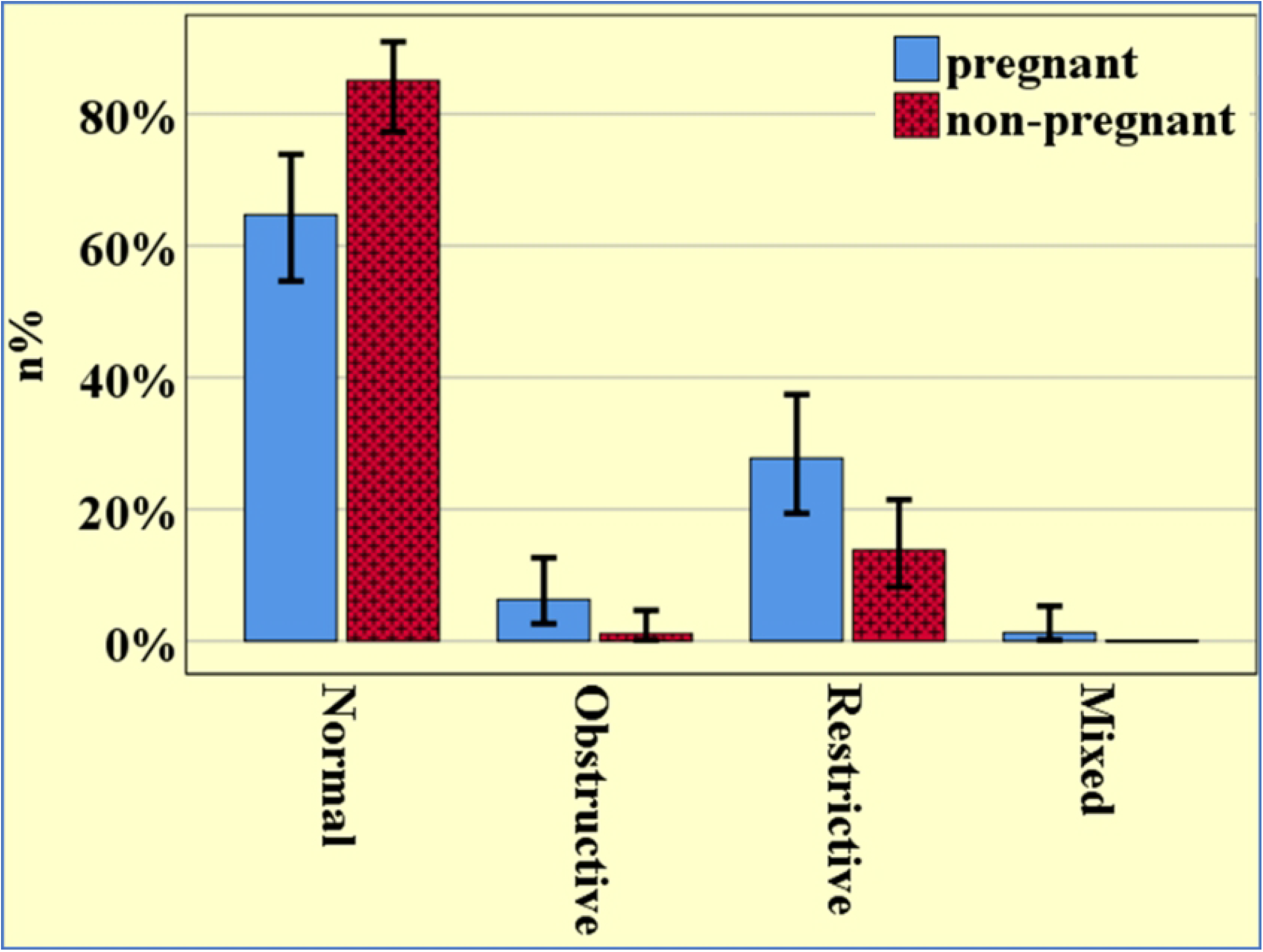
Spirometry pattern of lung function of pregnant and non-pregnant participants (n=196, Error bar=95% CI)

The spirometry performance quality grades were associated with body weight (p=0.015), height (p=0.018) and gestational age (p=0.014) but only body weight was significant after applying the logistic regression model. The acceptability of spirometry performance was not associated with any of the factors under study.

### Spirometry patterns of lung function during pregnancy

About 71.7% (n=66) of pregnant participants had normal patterns, while it was 92.6% (n=104) among controls. The pregnant participants had 2.5 times the odds of having a restrictive pattern (p=0.021, 95% CI=1.1-5.7) and remained significant even after adjusting for spirometry performance quality. The patterns were not associated with any of the participants’ characteristics or performance quality.

## Discussion

We studied spirometry performance among pregnant in comparison to non-pregnant women and evaluated whether test performance capability and quality substantially affect results. Our findings revealed that despite suboptimal test performance quality in pregnant as compared to non-pregnant women, the test results still portrayed physiologically expected patterns in pregnancy. Interestingly, we found no association between test quality grade and lung function patterns in pregnant and non-pregnant women.

Being pregnant increased the odds of achieving lower spirometry performance quality grades and more likely to produce unacceptable sessions than non-pregnant participants. While most pregnant participants had only two acceptable manoeuvres with repeatability exceeding 200 ml, most of the non-pregnant participants had three or more with repeatability not exceeding 200 ml. Also, pregnant participants were more likely to have restrictive lung function patterns.

Performing acceptable spirometry requires understanding and correct execution of given instructions. Lower-quality grades could be due to impaired cognitive function, especially attention, memory, and execution reported during pregnancy (12). Alternatively, pregnant participants could have had divided attention as they were attending antenatal services, hence failure to comprehend instructions for the correct execution (13). Yet, the lower acceptability quality of spirometry performance in pregnancy could be a contributing factor to the lower volumes as compared to their non-pregnant counterparts (8) although the values are believed to remain within normal limits (9).

Body weight independently correlated with spirometry performance during pregnancy. Weight change is a marked cardiometabolic alteration occurring during pregnancy and corresponds with physical performance (14). Also, we didn’t find any correlation between body age, height, and gestational age with spirometry performance, but these have been important predictors of test values (15). Contrary to the other studies (16,17), no factor under study was associated with the overall acceptability. This analysis included participants of close age and context and hence might have had less heterogeneity to reveal the effect of the factors.

In the current analysis, spirometry was more likely to classify pregnant women as having restrictive lung function patterns, as has been documented previously (18). This can be attributed to pregnancy-induced ribcage mu restriction and diaphragmatic changes due to an alteration in fibre length and abdominal pressure (19). The spirometry classification of lung function pattern was independent of quality grades; thus, A-C grades can still be clinically acceptable during pregnancy. Further studies can examine if spirometry interpretation criteria can be re-evaluated to accommodate suboptimal performance during pregnancy such as exploring the potential of using grade D under certain conditions. Although disregarded, the potential of lowering quality control criteria has been evaluated previously (20).

Our comparisons could have been limited as we did not assess the intellectual abilities, which could have influenced comprehension and therefore execution of instructions for correct spirometry (6). Also, pregnant participants were recruited randomly, while non-pregnant controls were recruited consecutively, and only pregnant participants who booked their first antenatal visit in their first trimester were included in the study.

## Data Availability

Data are available up on reasonable request from the author

## Acknowledgement

We acknowledge Mr Elibariki Kapinda the laboratory technician from MUHAS who conducted spirometry.

## Conclusion

Despite the higher likelihood of pregnant individuals producing suboptimal spirometry test quality, the observed pattern classification remains consistent with physiological expectations. This suggests that spirometry is still a valid tool for assessing lung function in pregnancy. However, the suboptimal performance rates among pregnant individuals highlight the need to reevaluate the use of spirometry in this population to ensure optimal testing conditions and accurate interpretation of results.

